# Alignment of Community Benefit Spending and Initiatives to Improve Community Health: Is There Evidence of Progress?

**DOI:** 10.1101/2022.08.17.22278878

**Authors:** Abel Sapirstein, Arthi Rao, Lauren N. Steimle

## Abstract

**Objectives:** We sought to identify changes in non-profit hospitals’ community benefit spending from 2014 to 2019. Secondly, we wanted to find novel predictors of spending in the most recent available data.

**Methods:** For our longitudinal analysis, we used tax filing data for 1072 hospitals from 2014-2019 and time-based ANOVA. We gathered information about hospital characteristics, the social determinants of health, and hospital partnerships with local communities for 1192 hospitals for the year 2019. We employed multivariate regression to identify significant factors.

**Results:** Total community benefit spending rose from 8.1% in 2014 to 9.1% of operating expenses in 2019, driven by increased spending on patient care. There was no such increase in spending on activities targeted at improving community health. The presence and strength of partnerships between hospitals and their communities were associated with higher community benefit spending.

**Conclusions:** We found no evidence of dramatic shifts in community benefit spending from 2014 to 2019. Further, we identified partnerships for population health improvement as an effective, novel predictor of community benefit spending. Supporting partnerships between hospitals and communities may help facilitate strategic investments in community health improvement.

## Introduction

In 2019, non-profit hospitals spent an estimated $81-110 billion on community benefits. ^1,2^ Community benefits include subsidizing direct patient care as well as activities that address social determinants of health (SDOH), such as community-building (e.g., housing, economic development, and environmental improvements) and community health improvement services. Investments in addressing SDOH provide an effective tool for improving health equity since such interventions target multiple modifiable risk factors for chronic conditions such as obesity, cancer, asthma, cardiovascular disease, and diabetes.^3–6^ These SDOH factors can also be strong predictors of preventable hospitalizations^7^ and emergency department utilization. ^8^ Thus, because more than 60% of hospitals in the US are non-profits, community benefit spending presents a significant resource to address upstream SDOH factors and associated health disparities. ^9^

Prior to the passage of the Affordable Care Act (ACA), most community benefit spending went to direct patient care, as opposed to investments in upstream factors influencing SDOH. ^10^ To better align community benefit spending with communities’ health needs, the ACA mandated additional requirements such as requiring that non-profit hospitals conduct Community Health Needs Assessments (CHNAs) to identify SDOH needs and make plans to address them. Despite this, community benefit spending had remained dominated by direct patient care benefits in the years immediately after the passage of the ACA. Young et al. reported that, from 2010 to 2014, overall spending on community benefits steadily increased from 7.6 % to 8.1% of total hospital expenses, and their observed total increase was driven by increased spending on direct patient care. ^11^ Only a small fraction of community benefit spending went to community health improvement initiatives and community building activities targeting SDOH factors.

Establishing which factors are associated with increased community benefit spending, particularly spending not on direct patient care, is of interest since such factors might refine policy for effective community investment. Past predictors of a hospital’s community benefit spending include hospital characteristics and the composition and needs of the communities that it services. ^11,12^ However, while increased funding in community health improvement initiatives is associated with fewer readmissions and preventable hospitalizations on a national scale, ^13^ neither community health improvement initiatives nor community-building activities appear to be responsive to community needs. ^12,14,15^ External partnerships and collaborations may be another determinant in community benefit spending. Previous research associated coordination between local health departments and hospitals in the CHNA and following implementation with increased spending on community health improvement initiatives. ^16,17^ However there has not been a study on the impact of partnerships for public health improvement beyond those with local health departments.

Previous studies concluded more time was needed for the ACA provisions for community benefits to change the distribution of spending to improve population health and better address SDOH proactively. ^11,15^ A decade removed from the initial implementation of the ACA, we present an updated analysis of community benefit spending. We evaluated distributional changes in community benefit spending between 2014 and 2019 and the extent to which institutional characteristics and contextual factors were predictive of community benefit spending. Further, we extend the relationship between hospital community benefit spending and the presence of partnerships beyond local health departments to show that the strengths of partnerships are among the strongest predictors of community benefit spending.

## Methods

Our study had two main objectives. First, we sought to identify changes in community benefit spending from 2014-2019. Second, we sought to identify predictors of community benefit spending during 2019. We constructed two different datasets for our analyses— one longitudinal and one cross-sectional. The longitudinal dataset allowed us to analyze changes in community benefit spending. To construct the longitudinal dataset, we scraped hospital financial data from Schedule H of IRS form 990 for the 1072 hospitals that had returns available for every year from 2014 to 2019. We constructed the cross-sectional dataset to analyze predictors of spending. To construct the cross-sectional dataset, we linked financial information about community benefit spending with hospital characteristics, community-level SDOH, and partnerships and connectedness for the fiscal year (FY) 2019 for 1192 hospitals. Non-profit hospitals report detailed community benefit expenditures across ten broad categories as well as general financial well-being on the IRS forms (Appendix C). Descriptive data about hospital characteristics, including details about what partnerships for public health improvement (PPHI) hospitals formed, were obtained from the 2019 American Hospital Association (AHA)’s annual survey. We converted PPHI information into an ordinal variable denoting the strength of the partnership. The four ordered levels of these partnerships corresponded to failure to report, no partnerships, informal collaborations, and formal partnerships. Lastly, 2019 census tract-level SDOH data was extracted from the Census Bureau’s American Communities Survey and were also compiled into a modified Community Needs Index (CNI), which combines independent SDOH factors into a composite measure to indicate healthcare barriers, using the methodology developed by Dignity Health. ^18,19^ Census tract-level SDOH and CNI data were aggregated up to the hospital service areas (HSA) as defined by the Dartmouth Atlas of Health Care (http://www.dartmouthatlas.org/) as these are more representative of a hospital’s healthcare market or community. To summarize, each record in the data table contained the hospital’s community benefit expenditures, hospital characteristics, aggregate SDOH characteristics of its service area, the CNI, and details about its PPHI for the year 2019. All data, except AHA survey data, were gathered from publicly available sources (see Appendix C).

### Key Variables and Measures

We profiled the level and type of hospital community benefit spending reported on the IRS Form 990 Schedule H forms to identify changes in spending and predictors of spending. We defined five different subtypes of community benefit spending) based on the categorization used by Young et al. ^11^: total community benefit spending (TCB), direct patient care spending (PCS), Medicaid shortfall spending (MS), community health improvement spending (CHI), and community building activity spending (CBA) (see Appendix A). To control for the correlation between each spending subtype and overall hospital expenses, all regressions and further analyses were conducted after normalization by total hospital expenses. Normalization by total hospital expenses also provided a way to account for inflation in healthcare spending.

To meet our second objective, we constructed three domains of predictors: hospital characteristics, SDOH, and connectedness and partnerships. Hospital characteristics contained information about which accreditations and programs a hospital had as well as details about how the hospital was classified by the Centers for Medicare and Medicaid Services. SDOH included the individual factors used to calculate CNI. ^18^ Lastly, connectedness and partnerships consisted of information about the presence of clinically-integrated networks, medical homes, and partnerships for public health improvement as defined by AHA survey data. PPHI were formed with 11 different potential partners, including local health departments, schools, health insurance companies, and healthcare providers outside of the hospital’s system. The components of each domain are listed in Table 1 with further details available in Appendix B.

**Table 1:** Our 11 sets of regressors across the 5 profiled spending subtypes. R^2^ reported as percentages of total explained variance and significance of models is denoted as * for p < 0.05 and ** for p < 0.01. Sets of regressors represent the permutation across all possible combinations of regressor blocks (hospital characteristics, SDOH, and community partnerships and collaboration). While none of the regressions were able to explain greater than 22% of the variance, the addition of information about partnerships and collaboration doubled the explained variance in total community benefits, patient care benefits, and MS. Further, we noted that SDOH had the greatest predictive capacity for community health improvement and community building activities. HC: hospital characteristics, SDOH: Social Determinants of Health, C+P: connectedness and partnerships

| Regressor Domains |  |  |  | Spending Subtype<br>R<sup>2</sup> % (Adj R<sup>2</sup> %) |  |  |  |
| --- | --- | --- | --- | --- | --- | --- | --- |
| Hospital<br>Chars | SDOH | C+P | Total<br>Community<br>Benefit | Patient Care<br>Benefits | Medicaid<br>Shortfall | Community<br>Health<br>Improvement | Community<br>Building<br>Activities |
| X | Indiv | X | 21.5 ( 17.3 ) ** | 21.6 ( 17.4 ) ** | 24.0 ( 20 ) ** | 8.4 ( 3.5 ) ** | 6.7 ( 1.8 ) |
| - | Indiv | X | 16.7 ( 14 ) ** | 15.9 ( 13.1 ) ** | 19.1 ( 16.5 ) ** | 6.0 ( 2.9 ) ** | 5.9 ( 2.8 ) ** |
| X | CNI | X | 14.7 ( 11 ) ** | 15.9 ( 12.3 ) ** | 14.6 ( 11 ) ** | 6.0 ( 1.9 ) * | 5.7 ( 1.6 ) |
| X | - | X | 14.0 ( 10.5 ) ** | 15.3 ( 11.7 ) ** | 14.5 ( 10.9 ) ** | 5.7 ( 1.7 ) | 5.4 ( 1.5 ) |
| - | CNI | X | 8.8 ( 6.7 ) ** | 9.1 ( 7 ) ** | 8.3 ( 6.2 ) ** | 2.8 ( 0.6 ) | 5.0 ( 2.8 ) ** |
| X | Indiv | - | 8.1 ( 6.5 ) ** | 7.7 ( 6.1 ) ** | 11.7 ( 10.1 ) ** | 5.1 ( 3.4 ) ** | 1.5 ( -0.3 ) |
| - | - | X | 8.1 ( 6.1 ) ** | 8.7 ( 6.8 ) ** | 8.3 ( 6.3 ) ** | 2.4 ( 0.3 ) | 4.5 ( 2.4 ) ** |
| - | Indiv | - | 5.4 ( 4.8 ) ** | 4.2 ( 3.5 ) ** | 9.1 ( 8.5 ) ** | 3.1 ( 2.5 ) ** | 0.5 ( -0.1 ) |
| X | CNI | - | 3.8 ( 2.7 ) ** | 4.5 ( 3.4 ) ** | 4.1 ( 2.9 ) ** | 2.6 ( 1.4 ) ** | 1.0 ( -0.2 ) |
| X | - | - | 3.2 ( 2.1 ) ** | 3.8 ( 2.8 ) ** | 3.7 ( 2.7 ) ** | 2.5 ( 1.4 ) ** | 1.0 ( -0.1 ) |
| - | CNI | - | 0.7 ( 0.6 ) ** | 0.5 ( 0.4 ) * | 0.1 ( 0.1 ) | 0.1 ( 0 ) | 0.0 ( -0.1 ) |

### Analyses

In line with our first objective, we generated descriptive statistics about CBS from 2014-2019. We compared spending trends across these 5 years among each of the subtypes of CBS and used ANOVA to test for significant changes in spending patterns across years.

In line with our second objective, we identified significant predictors of each community benefit spending subtype using linear multivariate regression for FY2019. FY2019 was chosen as it was both the first year that the AHA survey contained information about connectedness and partnerships and the most recent year for which most hospitals’ tax data was available. We conducted five groups of regression analyses; each analysis corresponded to one type of spending: TCB, PCS, MS, CHI, and CBA. For each group of regression analyses, we conducted regressions using eleven combinations of independent variables that are categorized according to the three key domains previously identified as potentially relevant to community benefit spending: hospital characteristics, SDOH, and partnerships and connectedness. All individual variables identified as statistically significant predictors of hospital community benefit spending in previous work were included in our regression analyses. ^11,12^ Definitions of individual regressors used in each variable block (hospital characteristics, SDOH, partnerships and connectedness) are available in Appendix B.

Lastly, we also calculated descriptive statistics about how hospital characteristics are related to the strength with which hospitals form partnerships with their communities and local stakeholders.

## Results

### Changes in Community Benefit Spending 2014-2019

Average reported total community benefit spending was 8.1% of operational expenses in 2014. By 2019, average total TCB had risen to 9.1%, a significant increase (ANOVA, p < 0.005) (Figure 1). This increase was largely driven by increased expenditures on subsidized health services, a subtype of direct patient care spending, which rose from 1.4% to 2.1% of operational expenses from 2014 to 2019 (ANOVA, p < 0.005). No significant increase was noted in MS, charity care spending, or the combination of MS and charity care spending. Community building activities remained stationary and low – accounting for less than 0.05% of total hospital expenses across all years studied. Other hospital services— such as expenditures on research, professional education (not shown), and community health improvement— remained similarly low when compared to patient care spending (Figure 1).

**Figure 1:**
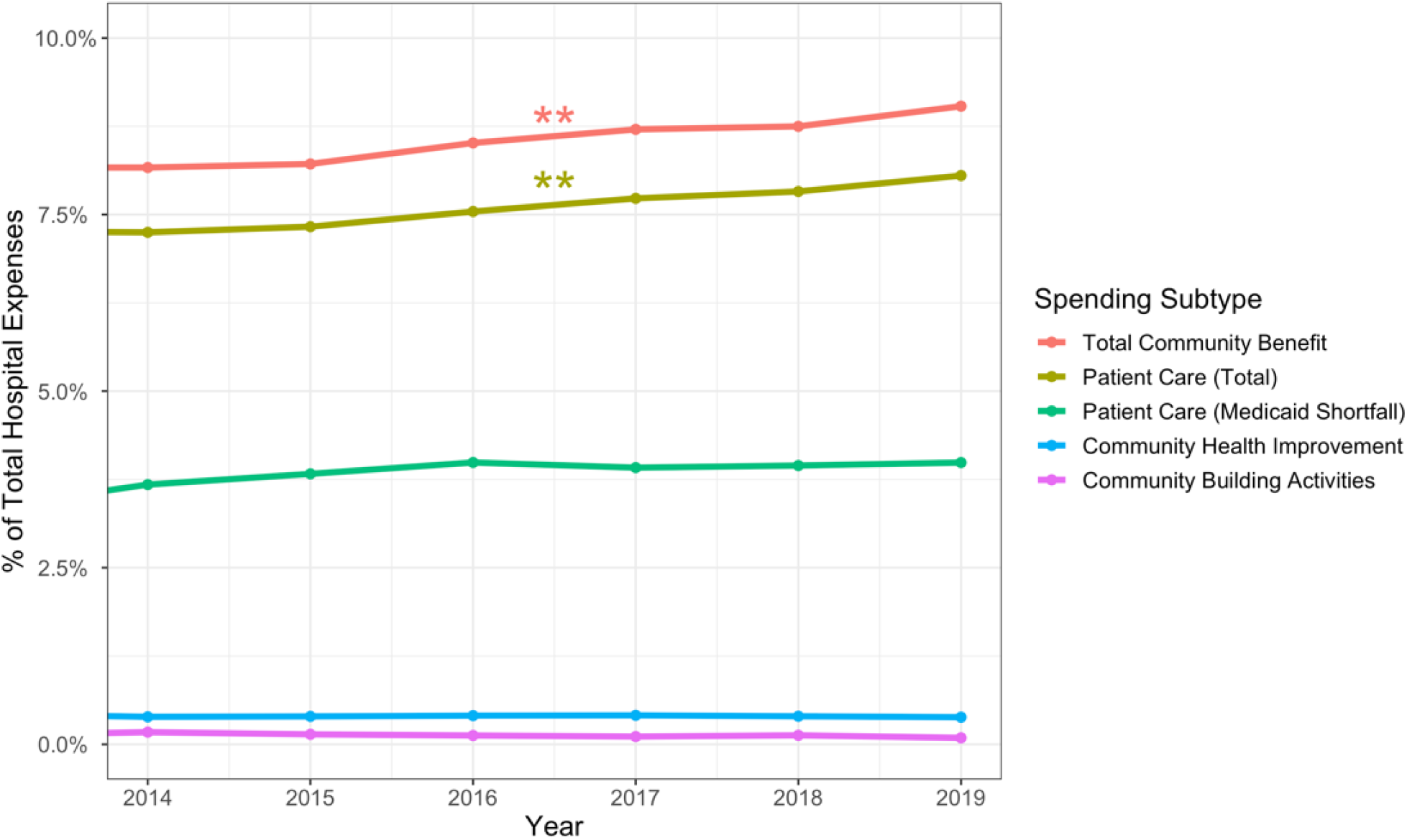
Total community benefit spending significantly increased from 2014-2019, from 8.1% of operational expenses to 9.1% of operational expenses. No increase was noted in community health improvement, community building activities, or Medicaid shortfall spending. Significant increases are denoted with **.

### Predictors of Community Benefit Spending

Table 2 summarizes which combinations of regressor blocks (hospital characteristics, SDOH, and connectedness and partnerships) explained significant variance across the 5 spending subtypes (TCB, PCS, MS, CHI, and CBA). We found that multivariate linear regression model built using a combination of hospital characteristics, single SDOH metrics, and connectedness and partnerships information formed the strongest set of predictors of TCB,PCS,MS. CBA were poorly predicted by all combinations of the hospital characteristics, SDOH, and partnerships and collaborations, with many combinations lacking any predictive power. Compared to other subtypes, predicting CBA is quite challenging given that a minority (48%) of hospitals reported any CBA, whereas a vast majority (98%) of hospitals reported PCS. Hospitals reporting no CBA had similar characteristics and were in communities with similar SDOH profiles to hospitals that reported community-building activities (Table 2).

**Table 2:** Results from regression with most explanatory power, significance levels are * < 0.05 and **< 0.01. In the instance of Community Building Activities, excluding hospital characteristics result in a better regression: as such they are excluded from the summary table. Regressors denoted with a ^+^ are ordinal regressors and the given coefficient corresponds to a one-level increase. Regressors denoted with a ^Ä^ are rate regressors and the given coefficient corresponds to a 1% increase in the regressor. All other regressors are binary variables, and the given coefficient denotes corresponds to the addition of the program/partnership. See Appendix for further details about SDOH

| Regressor | Total<br>Community<br>Benefit | Patient Care<br>Benefits | Medicaid<br>Shortfall | Community<br>Health<br>Improvement | Community<br>Building<br>Activities |
| --- | --- | --- | --- | --- | --- |
| R<sup>2</sup> (Adj R<sup>2</sup>) | 21.5 (17.3)** | 21.6 (17.4)** | 24.0 (20)** | 8.4 (3.5)** | 5.9 (2.8)** |
|  | Estimated Coefficient ± Standard Error |  |  |  |  |
| **Hospital Structural Characteristics** |  |  |  |  |  |
| Graduate Medical Education | -0.32 ± 0.62 | -0.14 ± 0.61 | -0.44 ± 0.41 | 0.06 ± 0.08 | - |
| Medical School Affiliation | **1.64 ± 0.59\*\*** | **1.64 ± 0.57\*\*** | **1.15 ± 0.39\*\*** | 0.07 ± 0.08 | - |
| DNV Accreditation | -0.52 ± 0.9 | 1.37 ± 0.88 | -0.36 ± 0.6 | -0.07 ± 0.12 | - |
| Teaching Hospital | **-2.03 ± 0.87\*** | **-2.17 ± 0.85\*** | **-1.61 ± 0.57\*\*** | 0.05 ± 0.12 | - |
| Medicare Certification | 0.69 ± 3.86 | -4.98 ± 3.78 | -5.07 ± 2.55* | -0.33 ± 0.52 | - |
| Joint Commission Accreditation | **-1.69 ± 0.68\*** | **-1.53 ± 0.66\*** | **-1.72 ± 0.45\*\*** | -0.13 ± 0.09 | - |
| Osteopathic Certification | **-2.4 ± 1.15\*** | **-2.71 ± 1.13\*** | **-2.95 ± 0.76\*\*** | -0.11 ± 0.16 | - |
| Catholic Church Operated | 0.85 ± 0.54 | 0.73 ± 0.53 | -0.31 ± 0.36 | 0.04 ± 0.07 | - |
| Critical Access Hospital | **-1.44 ± 0.66\*** | **-1.4 ± 0.64\*** | 0.01 ± 0.43 | -0.08 ± 0.09 | - |
| Rural Referral Center | **1.81 ± 0.7\*\*** | **1.44 ± 0.69\*** | **1.39 ± 0.46\*\*** | 0.18 ± 0.09 | - |
| Sole Community Provider Designation | 0.92 ± 0.95 | 0.49 ± 0.93 | 0.34 ± 0.63 | -0.06 ± 0.13 | - |
| Center for Improvement in Healthcare Quality | 1.04 ± 2.33 | 3.29 ± 2.29 | -0.77 ± 1.54 | 0.23 ± 0.31 | - |
| Accredited as Rehab Hospital | -0.45 ± 0.68 | 0.05 ± 0.67 | 0.09 ± 0.45 | -0.16 ± 0.09 | - |
| Cancer Program | 0.27 ± 0.53 | 0.2 ± 0.52 | -0.09 ± 0.35 | -0.11 ± 0.07 | - |
| **Social Determinants of Health** |  |  |  |  |  |
| Age 65+ in poverty<sup>Δ</sup> | 0.07 ± 0.08 | 0.01 ± 0.08 | 0.08 ± 0.05 | -0.02 ± 0.01 | -0.18 ± 0.14 |
| Minority composition<sup>Δ</sup> | 0.02 ± 0.02 | 0.01 ± 0.02 | **-0.06 ± 0.01\*\*** | 0.01 ± 0 | -0.03 ± 0.03 |
| Poor or no English fluency<sup>Δ</sup> | -0.17 ± 0.13 | -0.19 ± 0.13 | **0.22 ± 0.08\*** | -0.02 ± 0.02 | 0.08 ± 0.22 |
| No health insurance<sup>Δ</sup> | **-0.35 ± 0.06\*\*** | **-0.3 ± 0.06\*\*** | **-0.26 ± 0.04\*\*** | **-0.03 ± 0.01\*\*** | **0.38 ± 0.1\*\*** |
| Households w/ children in poverty<sup>Δ</sup> | -0.03 ± 0.1 | 0.04 ± 0.09 | **0.15 ± 0.06\*** | -0.02 ± 0.01 | -0.27 ± 0.17 |
| Female households w/ children in poverty<sup>Δ</sup> | 0.01 ± 0.04 | -0.01 ± 0.03 | **-0.05 ± 0.02\*** | 0.01 ± 0 | **0.12 ± 0.06\*** |
| Renting<sup>Δ</sup> | **0.07 ± 0.03\*** | 0.05 ± 0.03 | 0.03 ± 0.02 | 0.0 ± 0 | 0.04 ± 0.05 |
| Age 25+ w/o high school diploma<sup>Δ</sup> | **0.39 ± 0.08\*\*** | **0.37 ± 0.08\*\*** | **0.12 ± 0.06\*** | **0.03 ± 0.01\*** | 0.01 ± 0.14 |
| **Connectedness and Partnerships** |  |  |  |  |  |
| Medical Home Program - Hospital | 0.71 ± 0.55 | 0.2 ± 0.54 | **1.01 ± 0.36\*\*** | 0.04 ± 0.07 | 1.1 ± 0.95 |
| Medical Home Program - System | -0.04 ± 0.55 | 0.22 ± 0.54 | -0.71 ± 0.36 | 0.11 ± 0.07 | **2.31 ± 0.96\*** |
| Clinically Integrated Network - Hospital | -0.55 ± 0.55 | -0.32 ± 0.54 | -0.58 ± 0.36 | -0.02 ± 0.07 | -0.75 ± 0.96 |
| Clinically Integrated Network - System | -0.69 ± 0.59 | -0.52 ± 0.58 | **0.84 ± 0.39\*** | -0.17 ± 0.08* | 0.43 ± 1.02 |
| Patient and Family Advisory Council | **-1 ± 0.44\*** | **-1.07 ± 0.43\*** | -0.4 ± 0.29 | -0.03 ± 0.06 | -0.36 ± 0.76 |
| **Partnerships for Public Health Improvement** |  |  |  |  |  |
| Faith-based organizations<sup>+</sup> | 0.28 ± 0.44 | 0.29 ± 0.43 | -0.38 ± 0.29 | 0.11 ± 0.06 | 1.34 ± 0.69 |
| Health insurance companies<sup>+</sup> | **1.26 ± 0.35\*\*** | **1.25 ± 0.34\*\*** | **0.93 ± 0.23\*\*** | -0.03 ± 0.05 | 0.72 ± 0.9 |
| Healthcare providers outside your system<sup>+</sup> | -0.31 ± 0.41 | -0.21 ± 0.4 | **-0.76 ± 0.27\*\*** | 0.01 ± 0.05 | 1.05 ± 0.93 |
| Local business or chambers of commerce<sup>+</sup> | 0 ± 0.55 | 0.08 ± 0.54 | 0.52 ± 0.36 | -0.02 ± 0.07 | -0.28 ± 0.92 |
| Local or state human/social service orgs<sup>+</sup> | -0.55 ± 0.54 | -0.69 ± 0.53 | -0.03 ± 0.36 | 0.06 ± 0.07 | -0.93 ± 1 |
| Local or state public health orgs<sup>+</sup> | -0.61 ± 0.52 | -0.7 ± 0.51 | 0.1 ± 0.35 | 0.04 ± 0.07 | -0.31 ± 0.72 |
| National business<sup>+</sup> | 0.46 ± 0.4 | 0.37 ± 0.39 | 0.16 ± 0.26 | 0.02 ± 0.05 | -0.18 ± 0.6 |
| Non-profit organizations<sup>+</sup> | 0.22 ± 0.58 | 0.07 ± 0.56 | 0.45 ± 0.38 | -0.07 ± 0.08 | 0.72 ± 0.8 |
| Other<sup>+</sup> | 0.36 ± 0.45 | 0.53 ± 0.44 | 0.36 ± 0.3 | 0.03 ± 0.06 | -0.32 ± 0.95 |
| Other local or state government<sup>+</sup> | -0.66 ± 0.53 | -0.63 ± 0.52 | **-0.73 ± 0.35\*** | -0.01 ± 0.07 | -0.94 ± 0.69 |
| Schools<sup>+</sup> | -0.61 ± 0.46 | -0.63 ± 0.45 | **-1.08 ± 0.31\*\*** | -0.02 ± 0.06 | 0.53 ± 0.79 |

Although each spending subtype had a unique set of significant predictors, there was a shared core group that were significant across subtypes.(Table 2) Hospitals affiliated with medical schools spent on average 1.64% more of their total expenses on community benefit and PCS and 1.15% more of their total expenses on MS than their unaffiliated counterparts. Rural referral centers also spent more on TCB (1.81% more of total expenses), patient care spending (1.44 % more of total expenses), and MS (1.39% more of total expenses) than their unaffiliated counterparts. TCB, PCS and MS were all positively associated with insurance rates. A 1% increase in the insurance rate of an HSA was tied to roughly a 0.30% increase in TCB, PCS, and MS. This trend reversed for CBA, where the same 1% increase in insurance rate was tied to a 0.38% reduction in spending. Spending was also inversely associated with more education. A 1% increase in the percentage of HSA residents over age 25 without a high school diploma corresponded with a 0.39% increase in TCB, 0.37% increase in patient care spending, and 0.12% increase in MS. Hospitals that formed PPHI with health insurance companies tended to spend more on TCB, PCS, and MS than those that did not form such partnerships. This relationship was dependent on reporting and strength. On average, with every level of increase in collaboration, we saw a corresponding increase in spending of 1.26% of overall expenses for TCB, 1.25% of overall expenses for PCS, and 0.96% of overall expenses for MS.

The rate at which single mother households lived in poverty was associated with increased spending on CBA and MS but was not other subtypes. PPHI with providers outside of the hospital system and schools were associated with reduced MS but were not associated with any other subtypes.

Lastly, we present a comparative graph illustrating how different hospital types participate in PPHI (Figure 2). Detailed information on how different hospital types participate in individual kinds of PPHI is available in Appendix E. While the number of beds in a hospital was not a significant regressor for predicting any spending subtype, PPHI among large hospitals (400+ beds) were stronger and more abundant. Compared to non-critical access hospitals and hospitals that were not the sole health care providers in their communities, critical access hospitals and sole community providers had reduced partnership frequency and strength.

**Figure 2:**
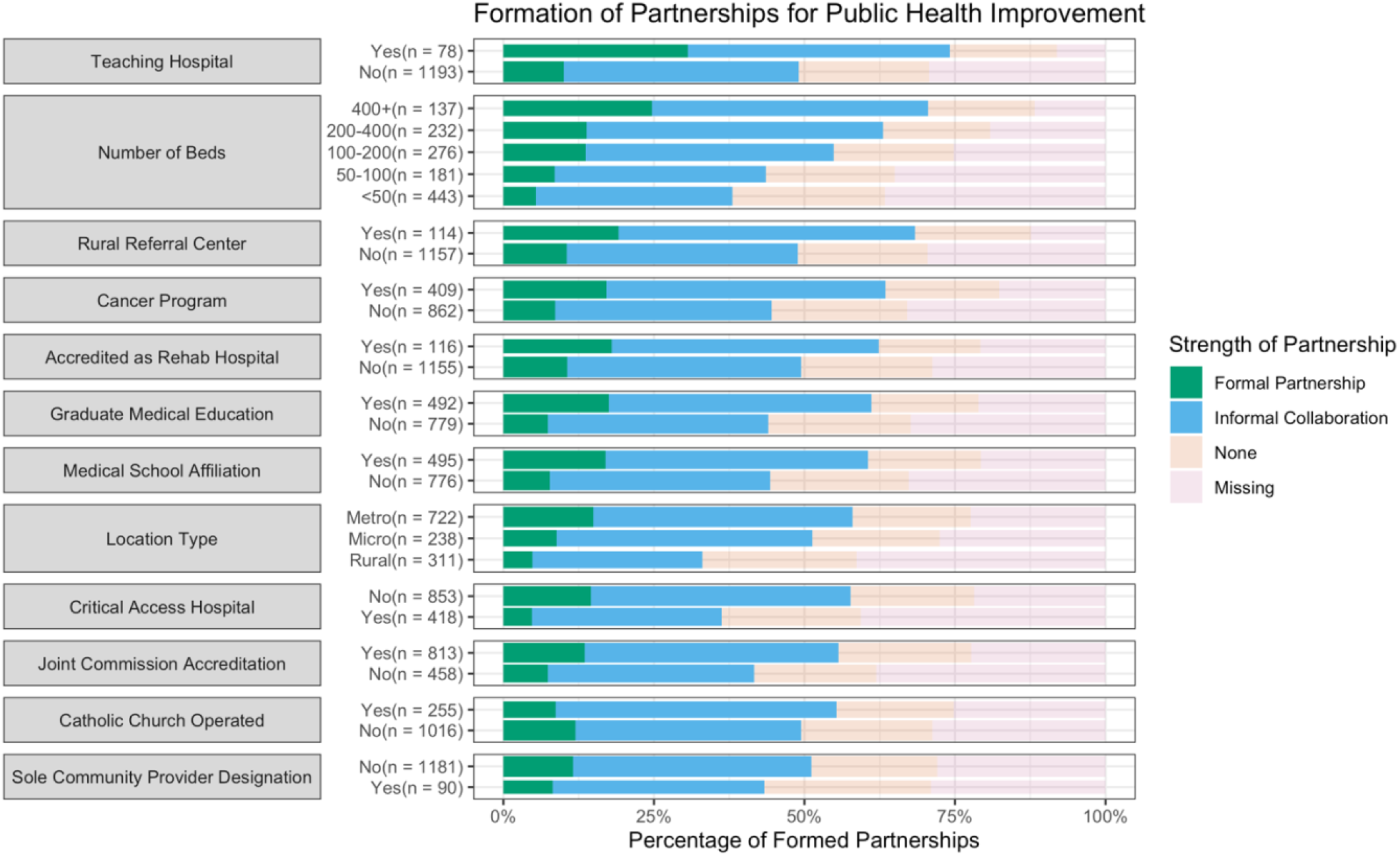
Prevalence of Partnerships for public health improvement, by hospital characteristics, colored by partnership strength. Large hospitals, teaching hospitals, and rural referral centers formed the greatest number of strong partnerships.

### Limitations of the Analysis

Our analysis is not without limitations. First, there are neither standard definitions of community benefit nor reporting requirements, implying that spending reported to the IRS does not provide the complete picture of community benefit expenditures. As such, hospitals lack a strong incentive to report all spending. Horwitz et al. (2020) estimated that health systems spent $2.5 billion on SDOH programs derived from other data sources, whereas IRS data for the same period only reports $902 million. ^20^. Given that most hospitals are non-profit hospitals and spend more on community benefit activities than for-profit hospitals, hospitals are under-reporting community benefit spending on tax filings. ^21^

Second, we only looked at hospitals that filed individual tax returns and omitted those that filed as part of hospital systems. We sought to profile the impact of community characteristics and partnerships in CBS, and aggregate hospital system spending information was unsuitable for this purpose. Additionally, consolidation and hospital closures have led to fewer hospitals filing individual tax returns, which is reflected in our smaller sample size in comparison to previous work.

## Discussion

Our analysis provides an update on the state of community benefit spending among non-profit hospitals in the US. We show a continued increase in TCB spending from 2014 to 2019, driven by an increase in spending on direct patient care, specifically subsidized patient care. This increase and its drivers are consistent with the increase and drivers observed from 2010-2014.^11^ Previous literature had suggested that additional time was needed for hospitals to make noticeable shifts towards addressing the health needs of their communities and align with the ACA’s policy goals. ^11^ With an additional 5 years of observation, we did not find evidence of a definitive shift in the distribution of TCB.

We sought to determine which factors were associated with increased hospital community benefit spending. Young et al. (2020) and Singh et al. (2015) previously identified the insurance status of the hospital’s likely patient population and hospitals’ teaching status as significant predictors of community benefit spending. We found these two factors remained significant predictors of some subtypes of TCB. ^11,12^ For example, hospitals in communities with many poor elderly residents were likely to suffer high burdens of MS. Peer hospitals located in areas with lower rates of elder poverty spent more on MS, despite similar reimbursement rates. Across these two regressors, magnitude and direction of coefficients also remained the same.

Community need, as measured by either SDOH indicators or the composite CNI, remained a poor predictor of CHI spending; hospitals located in high-need communities spend no more on CHI than their peers in low-need communities. This observation is consistent with previous findings by Singh et al. (2015). ^12^ Of all the community benefit spending subtypes, CHI is the least responsive to community. This is despite programs such as the CMS Hospital Readmission Reduction Program providing an external financial incentive to prioritize CHI. Hospitals in high-need communities serve patient populations at higher risk of readmission and thus are at greater risk of penalty in the CMS Hospital Readmission Reduction Program. Previous research has shown that CHI is associated with reduced readmission rates, showing evidence of direct benefits from investing more in CHI initiatives.^13^

We show PPHI between hospitals and their communities predict many subtypes of community benefit spending, with stronger partnerships yielding larger increases in community benefit spending. While previous work had established that partnerships between local health departments and hospitals resulted in greater TCB, our work is the first to extend this relationship beyond local health departments to other partners. We found that hospital partnerships with schools, insurance companies, or extramural healthcare providers, differed in TCB compared to their peers without such partnerships (Table 2). The directional impact of these PPHIs on TCB was not uniform. For example, PPHIs between hospitals and schools were found to be predictive of decreased MS, but not predictive of any overall change in PCS or TCB. PPHI with health insurance companies were associated with reductions in PCS. The explanatory power of PPHI was carried across rural vs. urban and bed-size differences. We also establish a strength-based relationship between PPHI and spending, with stronger partnerships showing a stronger effect. The PPHI domain was the strongest predictor among the three domains (hospital characteristics, SDOH, and PPHI) of hospital TCB, PCS, and MS and greatly improved the total variance explained by the models.

This finding expands the scope of previous work that has examined the relationship between hospital connectedness and community benefit spending. Singh and Young (2017) reported that local health department spending in the years 2009 (pre-ACA) and in 2013 (post-ACA) was not significantly associated with community SDOH spending by hospitals.^22^ However, a more recent analysis, showed that collaborations between hospitals and local health departments in performing CHNAs were significantly associated with more implementation activities and greater hospital spending on community health improvement initiatives in 2014.^16^ Our models also suggest that community partnerships are predictors of TCB but not specifically CBA or CHI spending, which are more directly related to improving SDOH than TCB. This trend is worth monitoring if such collaborations increase SDOH spending in communities through the CHNA mechanism.

Overall, there remains a clear need to address SDOH via widespread upstream interventions in areas such as housing, economic training, and patient education to improve community health.^23^ Despite this, we found that participation in — and reported funding for— community building activities remains low. While large hospitals were more likely to report community-building activities, less than 60% of even the largest hospitals (400+ beds) reported any spending on community building. This comes despite increasing evidence that investing in SDOH provides a good return on investment both in the form of reduced risk of readmission ^13^ and reduced morbidity and mortality. ^24^

We observed that the largest hospitals (400+ beds) formed partnerships with governmental organizations, with local health departments, with health insurers, and with external healthcare providers with greater abundance and strength than smaller hospitals (Figure 2). Teaching hospitals, which averaged 600 beds per hospital and spent more on TCB than their non-teaching peers, were even more likely to have reported the presence of the aforementioned partnerships (Figure 2). These partnerships could be potential future channels for SDOH funding.

While we were able to identify novel predictors of increased spending, assessing the true extent of spending to improve community health at a national scale remains difficult. Existing spending data is both coarse and underreported. Moreover, defining community need also presents a host of challenges. First, there is no consensus on the geographic area defining a hospital’s community. Though new methods, such as community detection algorithms, exist to solve this problem replicating them on a national scale is not feasible since geographic healthcare utilization data is not available nationally with sufficient detail and consistency.^25^ Further, even with accurately defined community areas, current SDOH data are not dynamic in time. Data sources such as the American Community Survey (ACS) are routinely used to quantify SDOH measures in communities. However, the ACS data are only aggregate 5-year averages and do not provide insight into variations within those periods, making it harder for healthcare systems to respond in a timely manner to community needs. Future work to improve the accuracy of and responsiveness to assessed community need may help hospitals better direct their investments and make it easier to demonstrate the benefits of such investments.

## Conclusions

We did not find evidence of dramatic resource shifts from 2014-2019 in hospital community benefit spending. A decade removed from the initial implementation of the ACA and 7 years removed from the original CHNA mandate, there are not clear signals that either policy resulted in a seismic shift in spending towards the stated US health policy goal of improved population health. We sought to identify what factors or hospital characteristics might be predictive of increased spending, both in overall spending and in promoting targeted spending on improving community health. Reported spending on community building activities and community health improvements remains non-responsive to community need. However, improved data reporting standards and mechanisms that help capture the wide variety of ways that hospitals might invest in their communities might change that narrative. We identified partnerships for population health improvement as an effective and novel predictor of community benefit spending. Supporting partnerships between hospitals and community stakeholders through more sophisticated data collection and integration mechanisms may help facilitate and direct strategic investments in SDOH program design, implementation, monitoring, and evaluation.

## Data Availability

All publicly available data can be gathered using to tools available at: https://github.com/asapirstein/cb_toolkit Additional data can be purchased from the American Hospital Assocation

## Acknowledgments

This research was supported by a Seed Grant awarded by the Office of the Executive Vice President for Research at Georgia Institute of Technology.

### Supplemental Materials

#### Appendix A

We define 5 spending subtypes of community benefit —these variables represent various logical aggregations of financial data available on each hospital’s form-990 tax return. All variables available in each 990 forms are listed below. Total Community Benefit spending is the sum of net expenditures in all fields marked **TCB**. Direct Patient Care Spending is the sum of net expenditures in all fields marked **PCS**. Medicaid Shortfall spending is the field marked **MS**. Community Health Improvement spending is the field marked **CHI**. Community Building Activity spending is the sum of net expenditures in all fields marked **CBA**.

| Item from 990 Form | Spending Subtype |  |  |  |  |
| --- | --- | --- | --- | --- | --- |
|  | TCB | PCS | MS | CHI | CBA |
| **Financial Assistance and Means Tested Government Programs** |  |  |  |  |  |
| Care at cost/ charity care | x | x |  |  |  |
| Medicaid | x | x | x |  |  |
| Other governmental means-based programs | x | x |  |  |  |
| **Other Benefits** |  |  |  |  |  |
| Community health improvement | x |  |  | x |  |
| Health professional education | x |  |  |  |  |
| Subsidized health services | x | x |  |  |  |
| Research | x |  |  |  |  |
| Cash and in-kind contributions | x |  |  |  |  |
| **Community Building Activities** |  |  |  |  |  |
| Physical improvements and health | x |  |  |  | x |
| Economic development | x |  |  |  | x |
| Community support | x |  |  |  | x |
| Environmental improvements | x |  |  |  | x |
| Leadership development | x |  |  |  | x |
| Coalition building | x |  |  |  | x |
| Community health improvement advocacy | x |  |  |  | x |
| Workforce development | x |  |  |  | x |
| Other | x |  |  |  | x |

#### Appendix B

As described above, we built multivariate regressions using three blocks of variables. Here we include details about these variables, data sources (either American Hospital Association (AHA) or American Communities Survey (ACS)) their encoding:

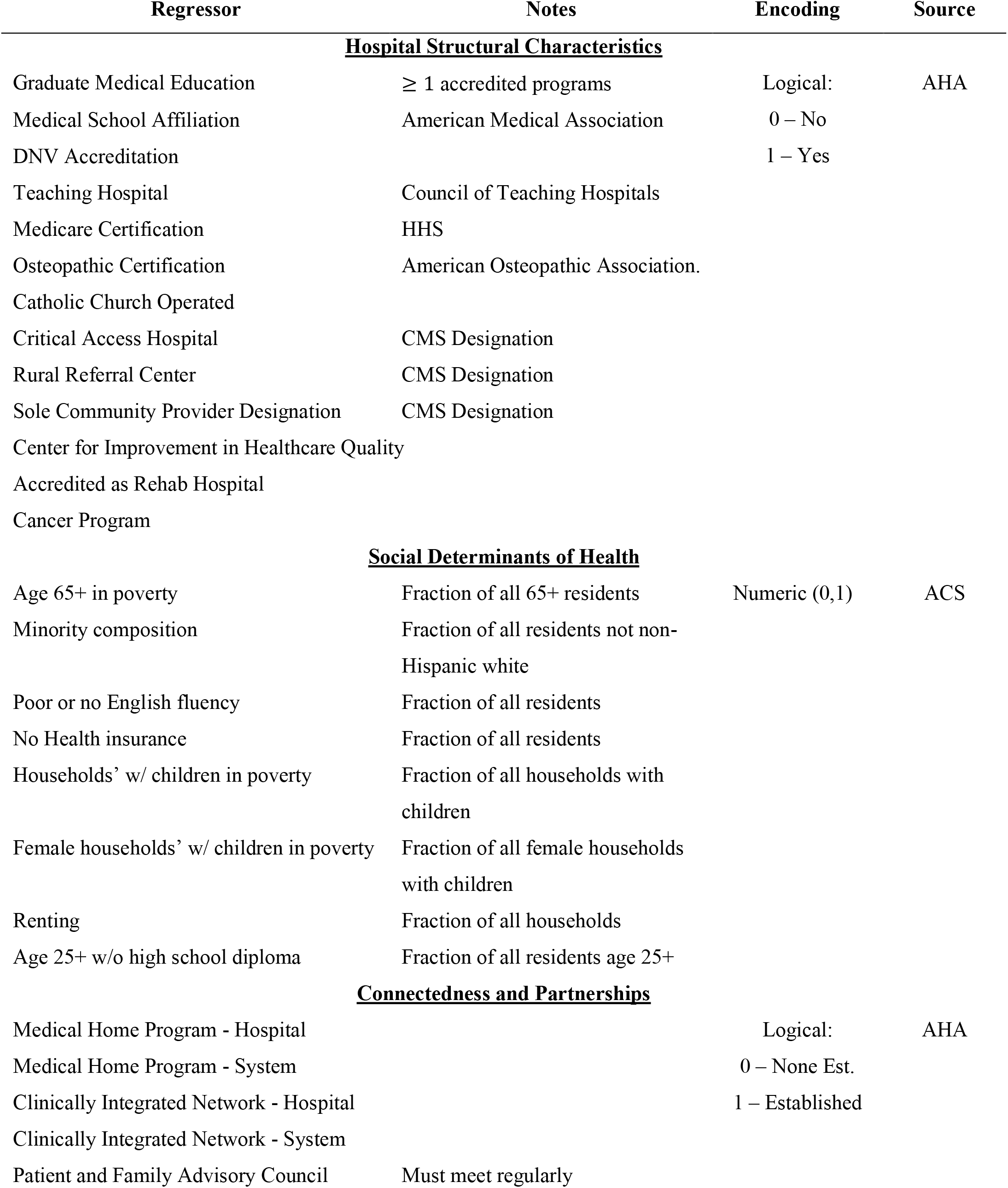
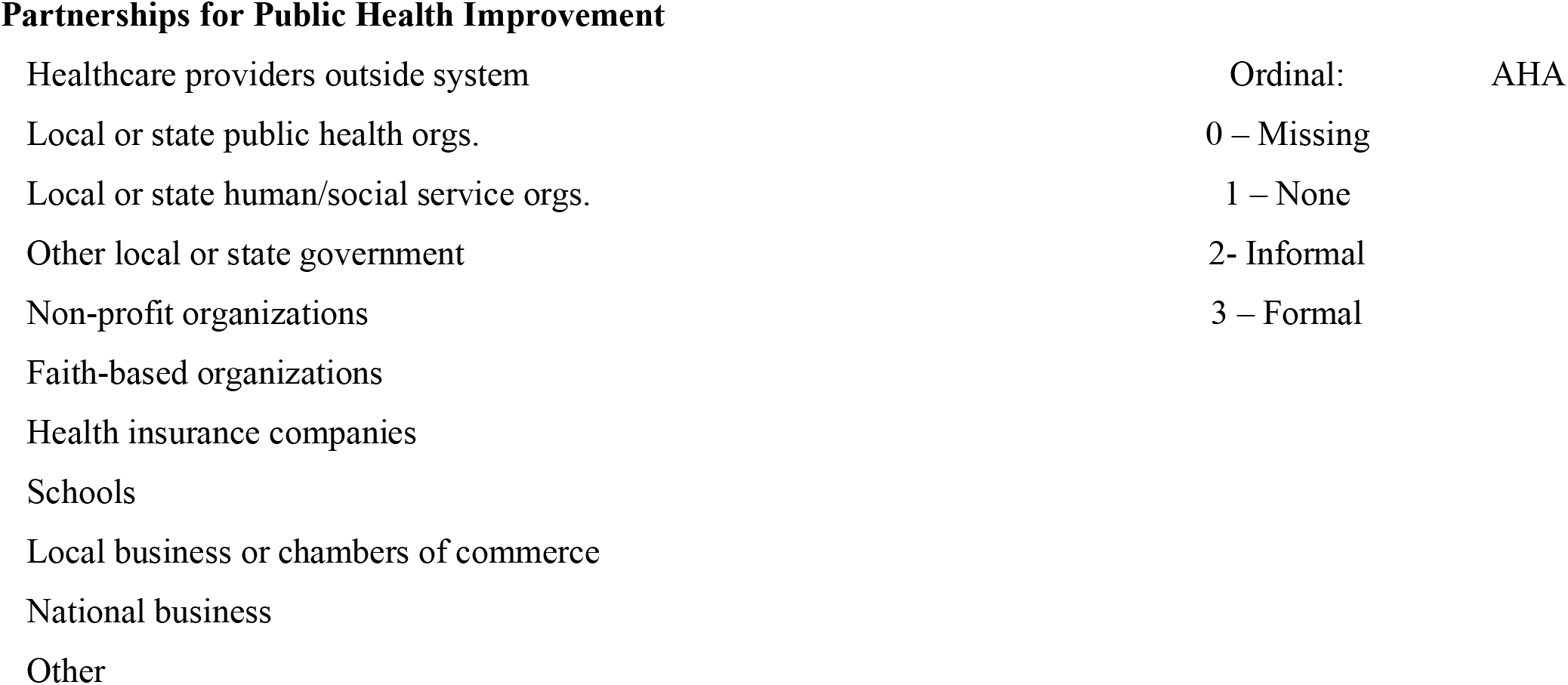

#### Appendix C

This appendix contains detailed information about data gathering and processing.

##### Financial data

Financial data was gathered by collecting machine-readable Form 990 Schedule H from the Registry of Open Data on AWS, which contains 990s for all non-profits in the US. We selected only those organizations filing Schedule Hs, which report hospital community benefit spending. Filing returns were linked to other hospital-level data by creating an Employer Identification Number (EIN) to CMS Certification Number (CCN) crosswalk. This crosswalk was generated by geocoding Form-990 filing addresses scraped from IRS data and hospital addresses provided on the CMS Provider of Service files. We then created a distance matrix from all EIN addresses to all CCN addresses and performed agglomerative clustering. For EIN locations more than a kilometer away from all CCN addresses, matching was performed manually, using Levenschtein distance to suggest likely matches based upon hospital name, state, and zip code. We found that such instances, where EIN location and CCN location differ, occurred most often when hospitals listed a corporate office on their tax return but a hospital location on their CCN. The remaining unmatched organizations were manually searched, if they were hospitals, we simply added their CCN to the crosswalk, otherwise, they were removed from further analysis. This EIN to CCN crosswalk included some hospitals that were not non-profits, these were removed by joining with purchased American Hospital Association data which included a list of non-profit CCNs.

##### SDOH Data

Social determinants of health (See Appendix B) were gathered from the American Communities Survey, using 5-year estimates with census tract resolutions. These census-tract level estimates were grouped by Hospital Service Area, with each census tract being assigned to the HSA which contained its centroid. The aggregate SDOH burden for a particular HSA was defined as the population-weighted average of that SDOH burden across all census tracts assigned to that HSA. CNI was approximated as an even weighting of 5 different barriers; an education barrier(percent of the population over the age of 25 without a high-school diploma); an income barrier composed of three evenly weighted components (elder poverty rate, child poverty rate, female household with children poverty rate); a cultural barrier with two evenly weighted components (minority rate and rate of poor/no English); an insurance barrier with two evenly weighted components (unemployment rate and percent of the population without health insurance); and a housing barrier (rental rate). This value was calculated for all HSAs, and then converted on a percentile on a 0-100 scale.

##### Code Availability

All code that was used to gather data from publicly available datasets is available at https://github.com/asapirstein/cb_toolkit. It is worth noting that this analysis was based on data gathered in December 2021. On December 31, 2021, the IRS announced that would no longer upload data to the AWS Open Data Initiative, as such no new financial data will be gathered by this scraper.

